# Neonatal EEG network activity associates with 2-year neurodevelopment after perinatal asphyxia

**DOI:** 10.64898/2026.05.26.26354098

**Authors:** Timo Syvälahti, Maksym Tokariev, Päivi Nevalainen, Anna Tuiskula, Marjo Metsäranta, Leena Haataja, Sampsa Vanhatalo, Anton Tokariev

## Abstract

**Background:** Prediction of long-term neurodevelopmental outcomes remains challenging after perinatal asphyxia. Here, we studied whether computational metrics of brain function derived from neonatal EEG are associated with long-term neurodevelopment in infants with perinatal asphyxia.

**Methods:** Total of 36 term-born infants with perinatal asphyxia with or without hypoxic-ischemic encephalopathy were studied with neonatal multichannel electroencephalography (EEG). We computed local EEG amplitudes and phase-amplitude coupling (PAC), as well as large-scale functional cortical networks estimated using amplitude-amplitude correlations (AAC) and phase-phase correlations (PPC). These EEG-derived markers were tested for associations with neurodevelopmental outcomes at two years, assessed using the Griffiths Scales of Child Development, 3rd edition (GMDS-III).

**Results:** EEG amplitudes showed positive associations with GMDS-III Foundations of Learning and General Development scores across most electrodes during quiet sleep, with the strongest effects observed at frontal and central regions (r = 0.44–0.66). PAC showed negative associations with the same scores mainly over parietal and temporal regions (r = −0.45 to −0.55). Cortical AAC networks demonstrated the most robust and widespread negative associations in all frequency bands during quiet sleep (r = −0.47 to −0.54), with 70–72% of connections significant in high delta frequency. In turn, PPC networks showed frequency-selective and more spatially constrained negative associations during quiet sleep (r = −0.48 to −0.53), involving 5–12% of the network.

**Conclusions:** Both local and network-based metrics in the newborn brain show significant association with neurodevelopmental outcome at 2 years after perinatal asphyxia.

## Introduction

Perinatal asphyxia associated with moderate or severe hypoxic–ischemic encephalopathy (HIE) is a major cause of neonatal brain injury and long-term neurodevelopmental impairment.^1–3^ Recent studies have shown that even perinatal asphyxia with no more than mild HIE may pose the infants at neurodevelopmental risks^4–6^. Moreover, it is increasingly accepted that the effects of perinatal asphyxia form a continuum rather than fall into discrete HIE categories^5,6^, prompting improved early identification of developmental risks to support immediate care and later follow-up.^7^ The currently used clinical criteria and neuroimaging provide valuable information, but they may not fully capture the functional spectrum of HIE consequences that is needed to reliably predict long-term development, especially after mild injury.

Electroencephalography (EEG) offers a noninvasive and widely available method to assess neonatal brain function. Most of the recent neonatal EEG literature has focused on the recovery of brain’s spontaneous activity (a.k.a. “background activity/pattern”) during the first 24-72 hours, typically using visual assessment of the raw signals^8–10^, amplitude integrated EEG trends^11,12^, or more modern interpretation algorithms^13–17^. It is now well established that recovery of the background activity during the first two days of life provides a particularly strong prediction of early and late clinical outcomes.^11,18–21^

However, it is less well understood how the more detailed characteristics of an apparently normalized background activity would link to later neurodevelopment. This gap in knowledge is striking, given the well-established link between all higher brain functions and the large-scale cortical network activity.^22–27^ Recent studies have demonstrated that neonatal multichannel EEG can be used efficiently for an objective, computational assessment of cortical functions both locally (amplitude or phase–amplitude coupling (PAC)) and globally, across brain-wide cortical networks (phase-phase correlation (PPC) or amplitude-amplitude correlation, (AAC)).^25,28–30^ These metrics are shown to link to a wide range of early life conditions, such as cortical maturation^25^, intrauterine drug exposures^31–33^, prematurity^34^, and exposure to early neurological insults^35,36^.

Importantly, our previous work showed that these same neonatal EEG network metrics exhibit graded alterations across the clinical continuum of perinatal asphyxia and HIE, and correlate with early clinical recovery after birth.^35^ Furthermore, the most recent publication with the same cohort showed that children exposed to perinatal asphyxia, even without clinical HIE, have persistent alterations in local cortical activity and large-scale cortical networks at two years of age.^37^ These findings suggest that EEG-based cortical network metrics may capture both early effects of perinatal asphyxia and long-term changes in cortical network activity. However, whether neonatal EEG network metrics are associated with later neurodevelopmental outcomes remains unstudied.

Here, we set out to study how the measures of local cortical function and large-scale networks associate with longer-term neurodevelopmental outcomes in infants with varying degrees of HIE after perinatal asphyxia. We computed these metrics from neonatal multichannel EEG recordings after perinatal asphyxia with or without HIE, and used regression analyses to test their associations with neurodevelopmental outcomes assessed at 2 years of age.

## Materials and methods

The overall study design and analytical flow is depicted in Figure 1 and described in detail in the following sections. In brief, we assessed four key EEG metrics across all neonates and sleep stages: (i) Frequency-specific amplitudes represented the overall oscillatory activity within each frequency band of interest. (ii) Phase-amplitude coupling (PAC) was computed to examine local cross-frequency interactions, which are indicative of intracortical network dynamics.^38^ (iii) Phase-phase (PPC) and (iv) amplitude-amplitude correlation (AAC) were calculated to evaluate connectivity-based interactions between cortical regions, as reflected by intrinsic connectivity modes.^22,23,26,39^ These metrics were compared with GMDS-III clinical assessments at age two years. A more detailed explanation of each metric is provided in the corresponding sections below.

**Figure 1.**
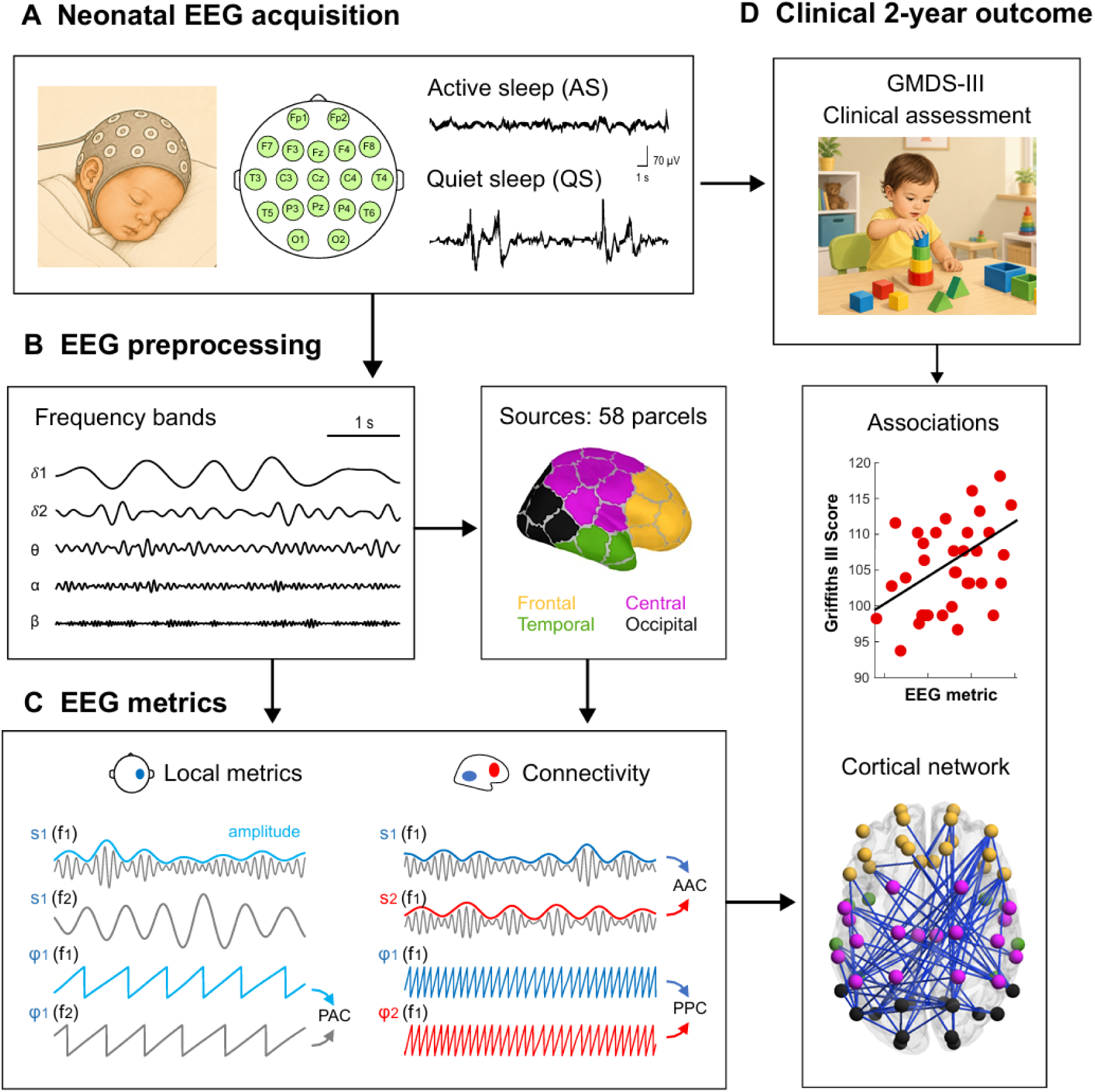
Overview of the study pipeline. (A) Multichannel EEG recordings were collected from neonates during daytime sleep and classified into active sleep (AS) and quiet sleep (QS) states. (B) Artifact-free 3-minute epochs were visually selected from each sleep states, filtered into five frequency bands of interest, and source reconstructed into 58 cortical parcel signals. (C) Four different EEG-based metrics were derived: local amplitudes and phase-amplitude coupling (PAC) were calculated at the scalp level for each electrode, while phase-phase correlation (PPC) and amplitude-amplitude correlation (AAC) networks were estimated for cortical parcels. (D) Metrics extracted from infant EEG were tested for associations with GMDS-III neurodevelopmental outcomes at 2 years using regression analyses.

### Study population

Our dataset included EEG data from a clinical cohort of patients with perinatal asphyxia (N = 36; gestational age ≥37 weeks). We recruited the neonates prospectively from the neonatal wards of the Helsinki University Hospital (Helsinki, Finland) and Jorvi Hospital (Espoo, Finland) in 9/2016 – 9/2020. Neonates were diagnosed with asphyxia by fulfilling at least one of the following criteria: 1-min Apgar score ≤ 6, umbilical arterial cord pH ≤ 7.10, need for assisted ventilation, or cardiopulmonary resuscitation after birth. Based on the modified Sarnat score^10^ and the amplitude-integrated electroencephalography (aEEG) during the first 24 hours, patients were classified as either having perinatal asphyxia without HIE (PA; N = 21), mild HIE (HIE1; N = 6), or moderate HIE (HIE2; N = 9). Standard clinical guidelines were used to decide on treatment with therapeutic hypothermia for each patient.^40^ EEGs were recorded at postnatal ages (PNAs) of 15.0 ± 10.4 days (PA: 18.7 ± 9.9 days, HIE1: 16.8 ± 10.3 days, HIE2: 5.1 ± 4.0 days; mean ± standard deviation) and postmenstrual ages (PMAs) of 42.7 ± 2.0 weeks (PA: 43.6 ± 1.4 weeks, HIE1: 42.1 ± 2.6 weeks, HIE2: 40.9 ± 1.6 weeks; mean ± standard deviation). The exclusion criteria for our cohort were lack of sufficient-quality neonatal EEG-recording, chromosomal abnormality, congenital anomaly, infection, concurrent neurological condition, severe HIE, or loss to follow-up. Clinical characteristics of this cohort have been described previously.^35,41^ The study design was approved by the Committee on Medical Research Ethics in the hospital district of Helsinki and Uusimaa (HUS/1331/2016), and guardians of the participating neonates gave their informed consent for the research.

### Neurodevelopmental follow-up

Neurodevelopmental outcome at 2 years was assessed by an experienced pediatrician (A.Tu.; blinded to the EEG results) using the GMDS-III. This standardized assessment evaluates five developmental domains, and a developmental quotient (DQ) score (mean 100, SD 15) is obtained for each subscale as well as the combined General Development score. For the present analyses, we focused on the Foundations of Learning, Personal-Social-Emotional, and General Development scores, as these domains were expected to reflect broader large-scale cortical network functions and overall neurodevelopment. We hypothesized that they would be most sensitive to early alterations in cortical networks.

Detailed cohort characteristics have been published previously.^41^ The mean Foundations of Learning DQ was 104.1 (SD 15.1; range 50–144), with four infants having score below −1 SD (<85), including one infant below −2 SD (<70). The mean Personal-Social-Emotional DQ was 104.3 (SD 6.1; range 91–118), with no infant scoring below −1 SD. The mean General Development DQ was 105.9 (SD 9.4; range 82–123), with five infants having score below −1 SD (<85), including two infants below −2 SD (<70). Mean age at GMDS-III assessment was 24.1 months (SD 0.7; range 22.7–26.1).

### EEG recordings

We recorded the 19-channel scalp EEGs (Fig. 1A) during daytime sleep with Waveguard caps with sintered Ag/AgCl electrodes (ANT-Neuro, Germany). We used either Eego EEG amplifier (ANT-Neuro, Germany) at a sampling rate of Fs = 500Hz or NicoletOne EEG system (Cardinal Healthcare/Natus) at a sampling rate of Fs = 2000 Hz to collect EEG data. Scalp electrodes were placed according to standard 10-20 scheme (see Fig. 1A). Electrode impedances were under 10 kOhm before recordings, and a trained expert monitored the signal quality continually.

### EEG visual review

All EEGs were primarily inspected in a clinical software (NicOne Reader, Natus). Trained specialist (T.S.) reviewed the data and selected artifact-free epochs of stable active (AS) and quiet sleep (QS) stages based on standard criteria (Fig. 1A).^42^ In AS, EEG showed continuous low voltage high frequency activity with irregular respiration pattern. In QS, EEG showed high-voltage slow frequency activity or tracé alternant (high-voltage bursts alternating with quiet periods) and respiration was regular. For each subject, 3 minutes of each sleep state (AS and QS) were selected for further analysis. One subject had an insufficient amount of good-quality AS data, and one subject had an insufficient amount of good-quality QS data.

### EEG preprocessing

EEG pre-processing was conducted using custom Matlab scripts (MATLAB R2022b, MathWorks, Natick, MA). The analysis utilized the same 19 common EEG channels (Fig.1A) across all participants in the study (Fp1, Fp2, F7, F3, Fz, F4, F8, T7, C3, Cz, C4, T8, P7, P3, Pz, P4, P8, O1, and O2). Initially, all EEG signals were band-pass filtered between 0.4 and 40 Hz, down-sampled to Fs = 100 Hz, and converted to average montage. For all analyses, signals were further filtered into five frequency bands (Fig. 1B): low delta (δ1: 0.4−1.5 Hz), high delta (δ2: 1.5−4 Hz), theta (θ: 4−8 Hz), alpha (α: 8−13 Hz), and beta (β: 13−22 Hz). Band-pass filtering was implementing using pairs of low-pass and high-pass Butterworth filters with the corresponding cut-off frequencies. Filtering was performed offline in a forward-reverse direction to prevent phase distortions and stopband attenuation in one direction was at least 15 dB. Scalp EEG signals were used to compute local amplitudes and PAC.

### Local EEG amplitudes

To estimate amplitude levels from band-pass-filtered EEG, we first applied the Hilbert transform to obtain the analytic representation of the signals. We then calculated the absolute values of the analytic signals to obtain amplitude envelopes. The mean values of these envelopes at each electrode were used as estimates of band-specific local amplitudes.

### Local phase-amplitude coupling (PAC)

The local cortical cross-frequency interactions were calculated with phase-amplitude coupling (PAC) metric, thought to reflect local connectivity between cortical layers (see Fig. 1C).^38^ PAC have been shown to correlate with maturation and vigilance states in neonates.^25,28,43^ In this study, we estimated PAC as previously described in detail^25,44^. In brief, from the same scalp electrode, we used the low-delta rhythm (0.4−1.5 Hz) as the ″nesting″ oscillation and higher-frequency rhythms (theta, alpha, and beta) as the ″nested″ oscillations. For the nested components, we computed amplitude envelopes using the Hilbert transform and filtered them with the same filter as the nesting component (0.4−1.5 Hz). PAC was estimated as phase locking value^45^ between the phase of the low-delta nesting component (φ_L_) and the phase of the amplitude envelope of the corresponding nested higher-frequency component (φ_H_):

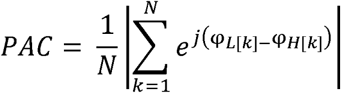

where *N* denotes the length of the time window, *k* is the sample index, *j* is the imaginary unit, and |·| indicates the absolute value.

### Source modeling

Before estimating functional networks, we first reconstructed cortical signals from scalp EEG recordings (Fig. 1B). To do so, we used a three-shell realistic infant head model^46^ comprising outer scalp, skull, and intracranial volume boundaries (2562 vertices per surface), with corresponding tissue conductivities of 0.43, 0.2, and 1.79 S/m.^47,48^ As the source space, we used the outer grey matter boundary approximated with 8014 vertices, each representing an electrical dipole oriented orthogonally to the surface. The forward solution was computed using the symmetric boundary element method (OpenMEEG^49^), and cortical source activity was estimated using dynamic statistical parametric mapping (dSPM^50^), as implemented in Brainstorm^51^. Further, cortical sources were clustered into N = 58 parcels according to an infant parcellation atlas^46^. Each parcel was further assigned to one of four anatomical categories based on its location (see Fig.1B): frontal (NC=C18), central (NC=C18), temporal (NC=C8), or occipital (NC=C14). Finally, parcel signals were computed as the weighted mean of all source signals belonging to the corresponding parcel.^51^

### Computation of cortical networks

Cortical functional networks were assessed using phase-phase correlations (PPC) and amplitude-amplitude correlations (AAC) between all pairs of parcel signals (see Fig. 1C).

These metrics capture complementary interaction modes in large-scale cortical networks: PPC quantifies rapid phase-synchronous coupling at sub-second time scales; whereas AAC reflects slower co-fluctuations in amplitude envelopes over multiple seconds across cortical regions.^22,23,26^ PPC was estimated using the debiased weighted phase lag index (wPLI):^52^

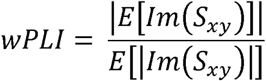

where E[·] denotes the expectation operator, Im(·) denotes the imaginary part, and S_xy_ denotes the cross-spectral density between the two time series x and y. AAC was calculated as the Pearson correlation between the amplitude envelopes of mutually orthogonalized signals (oCC):^53^

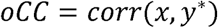

where y* denotes the version of signal y orthogonalized with respect to x, defined as y* = y − β·x, and β is the regression coefficient. Orthogonalization was performed in both directions: first y with respect to x, and then x with respect to y. The average of the two resulting oCC values was taken as the AAC estimate. These connectivity measures were selected as they are robust to volume conduction effects.^54,55^ The resulting connectivity matrices each comprised 58 × (58 − 1) / 2 = 1653 functional connections. However, our previous studies suggest that low-density EEG recording montages may be suboptimal for reliable reconstruction of whole-brain cortical activity in infants.^29,56^ Thus, we applied our previously developed simulation-based correction procedure to evaluate the 19-electrode head model used here and to exclude unreliable connections from further network analysis.^46^ This procedure generated a binary template in which reliable connections were marked as 1 and unreliable connections as 0. Applying this template to each connectivity matrix yielded 1128 reliable connections (the same across all subjects). PPC and AAC matrices were computed separately for five frequency bands and two sleep states, resulting in 10 matrices per neonate for each connectivity metric.

### Association with neurodevelopment

Associations between functional connectivity measures (PPC and AAC) and GMDS-III scores for Foundations of Learning, Personal-Social-Emotional, and General Development scores were assessed (see Fig. 1D) using separate network-based statistics (NBS) analyses^57^. For each analysis, a mass-univariate general linear model was fitted at each edge (functional connection) of the connectivity matrices, with GMDS-III scores used as regressors of interest. The design matrix included an intercept term, the corresponding developmental scores, and age at EEG acquisition. Postnatal age (PNA) and postmenstrual age (PMA) were significantly correlated and differed between clinical groups. To ensure that the observed associations were not driven by maturational changes^25,28^, we validated the results using two models: one with PNA (main text) and another with PMA (see Supplementary) as a covariate. Clinical severity (PA/HIE1/HIE2) was modeled using two binary indicator variables with the perinatal asphyxia (PA) group serving as the reference category.

The NBS analysis was implemented as follows. A t-statistic was computed for each unique pairwise connection in the connectivity matrices, and connections exceeding a primary threshold of t = 2.5 were selected. Connected components were then identified from sets of suprathreshold edges and the significance of each component was assessed based on its size.

Family-wise error (FWE)-corrected p-values were estimated using a nonparametric permutation procedure with 5000 iterations. For each permutation, subject labels in the design matrix were randomly permuted under the assumption of exchangeability, and a mass-univariate general linear model was fitted at each edge to compute t-statistics. The size of the largest connected component was recorded at each iteration, generating an empirical null distribution of maximal component size. Corrected p-values were computed as the proportion of permutations in which the maximal component size exceeded that observed in the original data. Components surviving correction at p_FWE_ < 0.05 were considered statistically significant.

To quantify the magnitude of effects within significant NBS components, edge-wise effect sizes were estimated post hoc. For each suprathreshold connection, t-statistics were converted to partial correlation coefficients using *r* = *t*/√*t*^2^ + *df*, where df represents the degrees of freedom in the model. The resulting r-values were averaged across all edges within each component to provide a descriptive summary of effect size. To quantitatively characterize the spatial scale of association networks, we computed network density (*k*) as the fraction of network connections showing significant associations. The involvement of distinct anatomical regions was assessed using ‘system load’, defined as the proportion of participating nodes from each anatomical category relative to full involvement of that category (nodes participating in all possible within- and between-category connections), and shown as the bars in Figures 4−5.

In addition to the NBS analyses, separate electrode-wise multiple linear regression analyses were performed to evaluate associations between local EEG measures (amplitudes and PAC) and GMDS-III developmental scores. The dependent variable was the corresponding GMDS-III score, and the predictor of interest was the electrode-specific EEG metric. Analyses were performed independently across all 19 electrodes for each frequency band and sleep state using the MATLAB *fitlm* function. We used the same covariates as in the NBS analysis and tested separate models including either PNA or PMA to assess robustness. For each model, t-statistics and p-values were extracted for the EEG predictor term, and partial correlation coefficients were derived from the corresponding t-values.

### Statistical analysis

We conducted all statistical analyses using MATLAB R2022b. Differences in GMDS-III domain scores between clinical severity groups were assessed with the Kruskal-Wallis test. Multiple comparisons were controlled using the Benjamini-Hochberg false discovery rate (FDR) correction procedure, with pFDR < 0.05 considered statistically significant. Local metrics were corrected across 19 electrodes for each frequency band and sleep state. Other statistical analyses are described in the corresponding sections.

## Results

### Clinical severity and age at EEG recording

The 2-year GMDS-III scores showed no significant group level differences between infants with different HIE severity (PA, HIE1, HIE2; Kruskal–Wallis tests, χ² = 0.02–1.59; all p > 0.9; Supplementary Fig. 1). There were also no significant correlations between postnatal age (PNA) or postmenstrual age (PMA) at EEG recording and any of the 2-year GMDS-III scores (Spearman ρ = −0.11 to 0.083, all p > 0.5; Supplementary Figs. 2 and 3).

### Local EEG amplitudes

Local EEG amplitudes showed widespread positive associations with GMDS-III Foundations of Learning and General Development scores in quiet sleep (Fig. 2 & Supplementary Fig. 4). In the significant channels, partial correlations ranged from r = 0.44 to 0.66 (p = 0.013 to < 0.001, pFDR = 0.048 to < 0.001), with the strongest effects observed for General Development score in the theta band (peak r = 0.66, p < 0.001, pFDR < 0.001 at Fp2 channel) and for General Development score also in the delta bands (peak r = 0.65, p < 0.001, pFDR < 0.001 at C4 channel). Foundations of Learning score showed similar but slightly weaker associations, with peak effects r = 0.51 (p = 0.0031, pFDR = 0.015) in δ1 at Fp2 channel, r = 0.55 (p = 0.0011, pFDR = 0.0056) in δ2 at C4 channel, and r = 0.56 (p < 0.001, pFDR = 0.0044) in θ at Fp2 channel. Spatially, the effects were broadly distributed, most prominently in frontal and central regions. No significant associations were observed with Personal-Social-Emotional scores. In active sleep, associations were limited: only Fp2 channel showed a significant association with Foundations of Learning score in delta band (r = 0.45, p = 0.0093, pFDR = 0.046).

**Figure 2.**
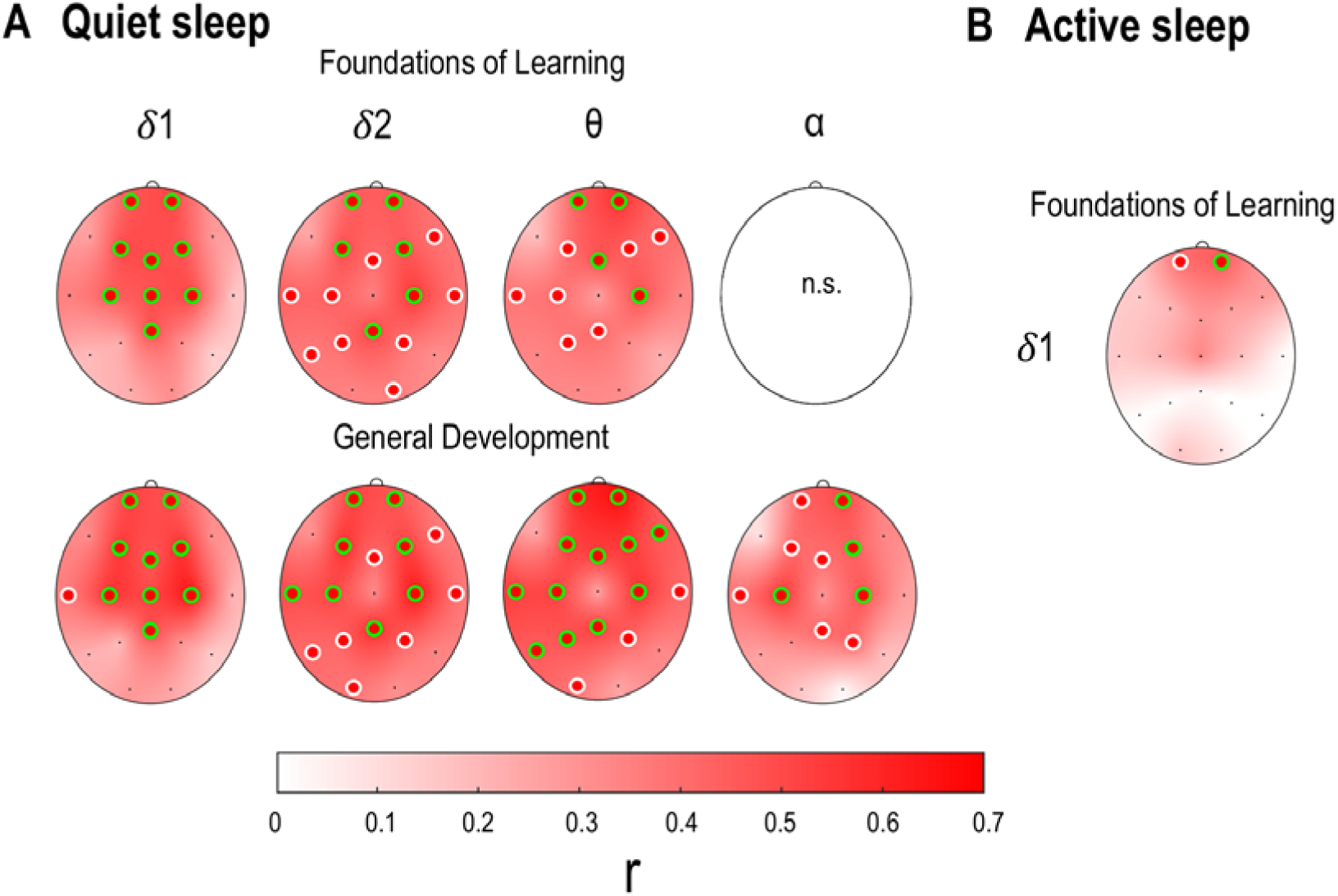
Local EEG amplitudes vs. 2-year neurodevelopmental outcomes. Scalp colormaps show associations between local EEG amplitudes and 2-year GMDS-III Foundations of Learning and General Development scores in quiet sleep (A) and active sleep (B) across the low delta (δ1), high delta (δ2), theta (θ), and alpha (α) frequency bands. Color indicates the strength of association, expressed as partial correlation (r) derived from the regression model. Circles indicate electrodes where amplitudes showed significant associations with GMDS-III scores (p < 0.05), while green circles denote electrodes that also passed FDR correction for multiple comparisons. N.s.: no significant association. Associations were mainly concentrated in frontal and central regions.

### Local phase-amplitude coupling (PAC)

Local PAC showed negative associations with GMDS-III outcomes (Fig. 3 & Supplementary Fig. 5). In quiet sleep, significant associations were found for both Foundations of Learning and General Development scores, with partial correlation coefficients ranging from r = −0.45 to −0.55 (p = 0.010–0.0011, pFDR = 0.0054–0.047). For Foundations of Learning score, peak effects reached r = −0.50 (p = 0.0038, pFDR = 0.018) in the θ band at P3 channel, r = −0.52 (p = 0.0022, pFDR = 0.011) in the α band at T5 channel, and r = −0.51 (p = 0.0031, pFDR = 0.015) in the β band at T5 channel. For General Development score, the strongest associations were seen in the β band, reaching r = −0.55 (p = 0.0011, pFDR = 0.0054) at T5 channel, while peak effects in the θ and α bands were r = −0.51 (p = 0.0029, pFDR = 0.014) and r = −0.51 (p = 0.0028, pFDR = 0.013), respectively, at P3 channel. Spatially, the effects were concentrated mainly over posterior parietal and temporal regions. No significant associations were observed with Personal-Social-Emotional scores. In active sleep, associations were more limited and only observed for General Development score, with peak effects of r = −0.48 (p = 0.0053, pFDR = 0.026) in the α band at Cz channel and r = −0.47 (p = 0.0064, pFDR = 0.031) in the β band at O2 channel.

**Figure 3.**
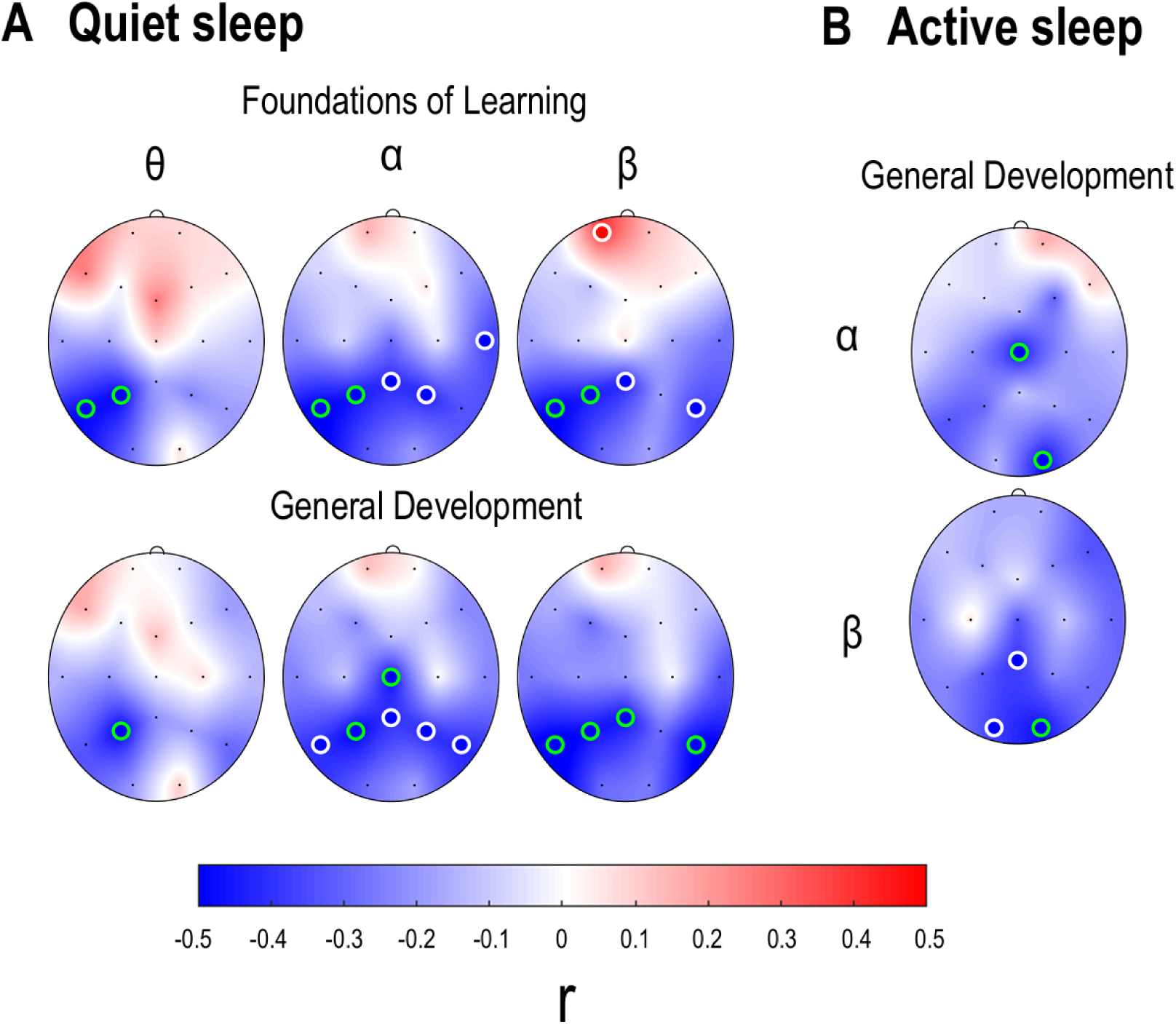
Local phase-amplitude coupling (PAC) vs. 2-year neurodevelopmental outcomes. Scalp colormaps show associations between local PAC and 2-year GMDS-III Foundations of Learning and General Development scores in quiet sleep (A) and active sleep (B) across the theta (θ), alpha (α), and beta (β) frequency bands. Color indicates the strength of association, expressed as partial correlation (r) derived from the regression model. Circles mark electrodes at which PAC was significantly associated with the corresponding GMDS-III scores (p < 0.05), with green circles indicating electrodes that also survived FDR correction across electrodes. Note, that significant negative associations were mainly concentrated in parietal regions.

### Cortical networks

PPC cortical networks showed negative associations with GMDS-III Foundations of Learning and General Development scores in quiet sleep (Fig. 4 & Supplementary Fig. 6). For Foundations of Learning score, significant networks were observed in the δ2, θ, and α bands, with effect sizes of r = −0.52 to −0.53 (pFWE = 0.0016–0.037), and k = 7–12% (k: % of connections showing significance). The patterns were spatially constrained and mostly proportionally distributed across different anatomical regions. However, temporal nodes showed slightly elevated involvement in the θ-band network, reaching 22% of their maximum possible participation. For General Development score, significant networks were found in the θ and α bands, both with r = −0.48 (pFWE = 0.003 and pFWE = 0.039, respectively), with k = 6% in the θ band and k = 5% in the α band. In active sleep, significant positive association was found for Foundations of Learning score in the θ band (r = 0.47, pFWE = 0.028, k = 6%). For Personal-Social-Emotional score, a mild positive association was observed only in the δ2 band after PMA adjustment (r = 0.46, pFWE = 0.03, k = 3%; Supplementary Fig. 6), but not after PNA adjustment.

**Figure 4.**
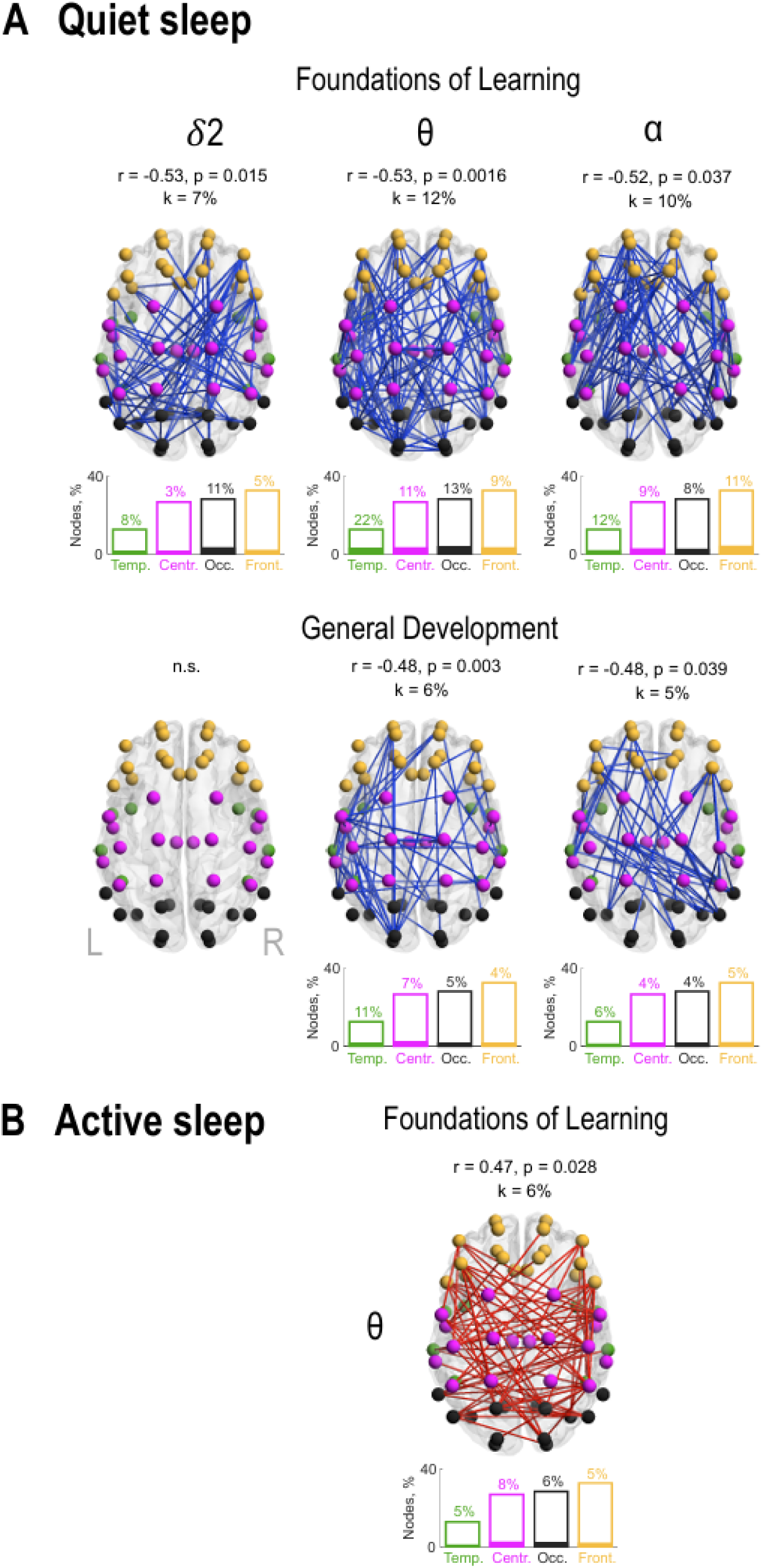
Cortical PPC networks vs. 2-year neurodevelopmental outcomes. Network plots show significant associations between large-scale phase–phase correlation (PPC) networks and GMDS-III Foundations of Learning and General Development scores in quiet sleep (A) and active sleep (B). Frequency bands are shown above each plot (δ2: high delta, θ: theta, α: alpha). Blue networks indicate negative associations and red indicate positive associations. Node colors highlight distinct anatomical regions: temporal = green, central = magenta, occipital = black, and frontal = orange. Bar height indicates the maximum possible involvement of nodes from each anatomical region, while the filled portion shows their observed involvement in specific networks. Partial correlation coefficients (r), derived from the regression models, corresponding family-wise error-corrected p-values, and network densities (k) are shown above each network. N.s.: no significant association.

AAC cortical networks showed widespread negative associations with GMDS-III outcomes in quiet sleep (Fig. 5 & Supplementary Fig. 7). For Foundations of Learning score, significant networks were observed across all frequency bands, with effect sizes ranging from r = −0.48 to −0.54 (pFWE = 0.005 to 0.001) and total network involvement of k = 32–72%. The strongest effect was seen in the α band (r = −0.54, pFWE = 0.001, k = 59%), while the most spatially extensive network was observed in δ2 band (r = −0.52, pFWE = 0.0006, k = 72%) with high involvement across all regions. For General Development score, associations were also found across all frequencies (r = −0.47 to −0.51, pFWE < 0.001), with the strongest effect in δ2 band (r = −0.51, pFWE < 0.0001, k = 70%). For Personal-Social-Emotional score, less widespread associations were found in the δ1 (r = −0.46, pFWE = 0.022, k = 10%) and δ2 bands (r = −0.45, pFWE = 0.048, k = 4%). In active sleep, findings were more limited, with significant associations for Foundations of Learning score in the α (r = −0.49, pFWE = 0.012, k = 20%) and β bands (r = −0.50, pFWE = 0.0086, k = 16%), and for General Development score in the β band (r = −0.47, pFWE = 0.042, k = 6%).

**Figure 5.**
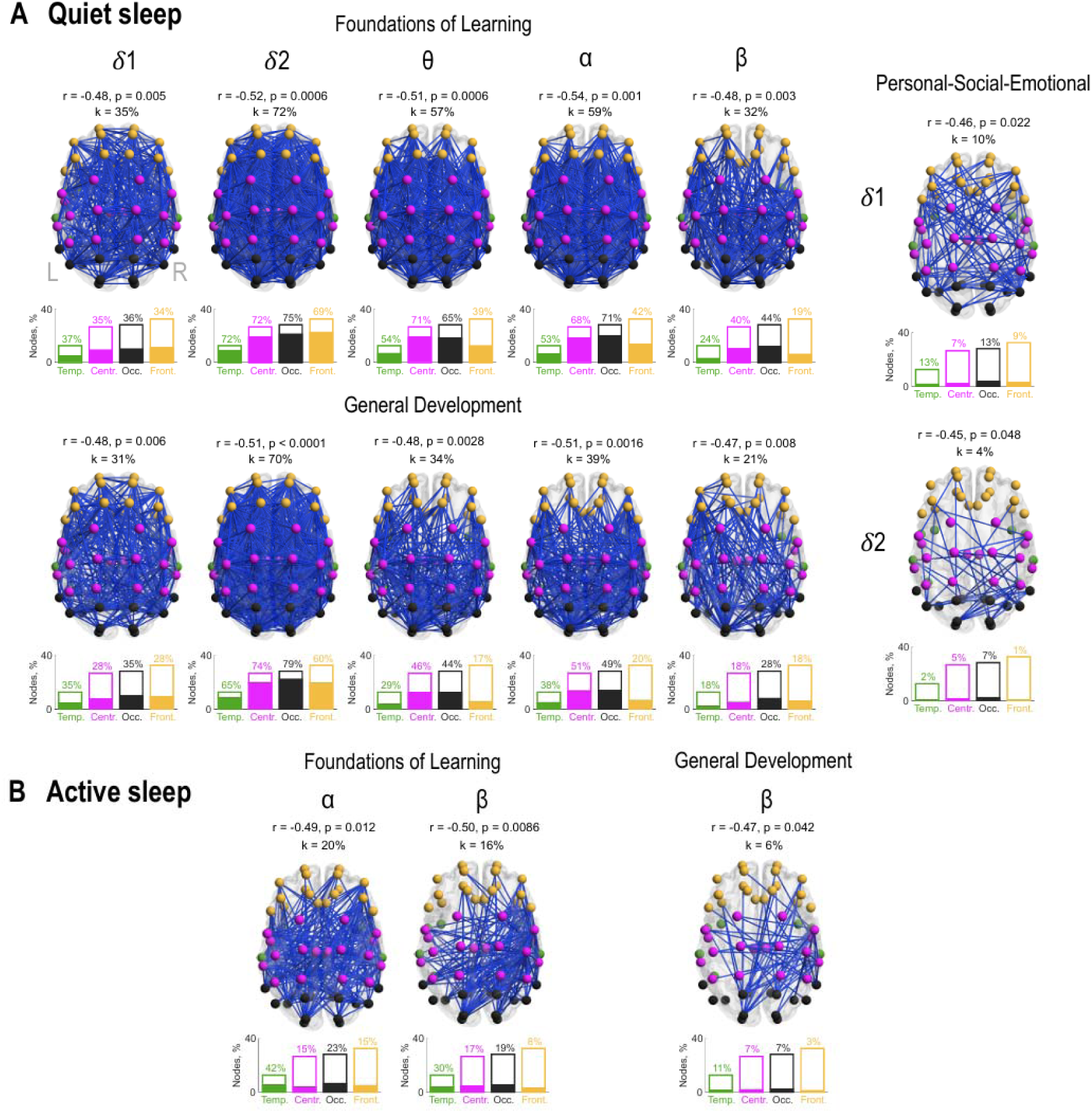
Cortical AAC networks vs. 2-year neurodevelopmental outcomes. Network plots show significant associations between large-scale amplitude–amplitude correlation (AAC) networks and GMDS-III Foundations of Learning, Personal-Social-Emotional, and General Development scores in quiet sleep (A) and active sleep (B). Frequency bands are shown above or next to each plot (1: low delta, 2: high delta, θ: theta, α: alpha, β: beta). Node colors indicate anatomical regions (temporal = green, central = magenta, occipital = black, and frontal = orange), and the bar plots below each network show the involvement of specific anatomical regions in the association network, indicated by the filled portions of the bars. Partial correlation coefficients (r), derived from the regression models, corresponding family-wise error-corrected p-values, and network densities (k) are shown above each network. Associations were observed across all areas, though strongest in the occipital region and in the high delta frequency band.

## Discussion

Our findings show that both local EEG metrics and large-scale cortical networks, analyzed from short sleep EEG recordings during the first weeks of life, are associated with neurodevelopmental outcomes at two years of age in infants with perinatal asphyxia. Prior studies correlating neonatal EEG to long-term developmental outcome have typically focused on a few pre-selected EEG metrics and/or cohorts with clearly abnormal outcomes.^15,17,58–61^ Here, we extend the previous literature by systematically characterizing association of different EEG network metrics and 2-year neurodevelopmental outcomes in a cohort where the majority of infants develop within normal range. These results also extend previous findings showing that the same EEG metrics correlate with neonatal clinical factors, such as the HIE severity and early clinical recovery.^35^ Together, these findings highlight the potential of neonatal EEG network analyses for an early recovery assessment and long-term prognostication.

Previous studies on outcome prediction after perinatal asphyxia with EEG or amplitude-integrated EEG have focused primarily on visual or automated EEG background features.^8,9,13,14,20,58–62^ It has been shown that visual^8,9^ or automated^13,61^ EEG background analysis, and the presence of seizures^62^ are predictive of adverse neurological outcome at two years. However, to our knowledge, there has been no prior studies linking EEG network activity to long-term outcomes in infants with perinatal asphyxia. Furthermore, earlier studies have typically used relatively extreme outcomes, such as death, severe neurodevelopmental impairment, or Bayley scores below −2 SD^14,59,60^. In contrast, our study extends prior literature by demonstrating that EEG network metrics are associated with neurodevelopmental variation even within the normal outcome range, suggesting that these measures may capture more subtle aspects of early brain function that are not reflected in conventional outcome classifications.

Earlier studies with similar EEG network metrics have reported that amplitude and coupling metrics, such as PAC^35,63,64^ and ASI^65^, are sensitive to perinatal insults^35,36^ and drug exposures in the neonatal period^32,33^. Prior neonatal EEG studies showed that functional brain networks dynamically change with maturation^25,28^, depend on sleep state^25^, and are affected by early-life adversities^32^. Importantly, PPC networks have been shown to link with neurodevelopmental outcomes at two years in preterm infants.^34^ The present findings build on this literature by demonstrating that variation in these EEG metrics in perinatal asphyxia is not only associated with current brain state, but also meaningfully related to developmental performance years later.

In our previous study using the same cohort, we found clear differences in EEG metrics across the clinical severity groups of perinatal asphyxia and hypoxic-ischemic encephalopathy (PA, HIE1, HIE2) in the neonatal period.^35,36,41,63,66,67^ However, the discrete perinatally assigned clinical HIE scores did not differentiate neurodevelopmental outcomes at two years of age (see Supplementary Fig. 1). This observation is in line with the emerging view that early discrete HIE grades are not able to fully reflect the spectrum of long-term developmental sequalae, particularly among infants who initially appear to develop within the normal range.^4,68,69^ Indeed, infants with HIE developing initially normally, may have difficulties in higher executive functioning and behavior even at school age.^70–72^ Thus, computational EEG network metrics may capture subtle functional brain alterations that are not reflected in categorical diagnoses, but which potentially carry predictive value for later development.

This study has some limitations. First, the sample size is relatively small, which limits statistical power, especially in the analysis of large-scale connectivity networks involving multiple frequency bands and sleep stages. The present findings should be considered primarily exploratory, and larger and more variable cohorts are needed to assess generalizability of these findings. Second, age-related variability is a potential confounder because many neonatal EEG metrics may change within weeks around term age^6,10,33^, and in our cohort differences in age was linked to clinical severity groups. We aimed to mitigate this uncertainty by having both postmenstrual and postnatal age at the time of EEG recording as covariates. Future studies with significantly larger cohorts are needed to allow more rigorous and balanced control of age and HIE grades.

In conclusion, our findings suggest that computational EEG-based metrics obtained during the neonatal period may serve as early functional biomarkers of both perinatal brain injury and later neurodevelopmental outcomes. These results highlight the potential of local EEG markers and network-based analyses to provide clinically meaningful information that complements traditional clinical assessments. Further studies are needed to confirm these associations and to evaluate their potential role in clinical decision-making and early intervention strategies.

## Data Availability

Original data consists of sensitive patient data that can not be shared. Anonymized association matrices and other derivatives are available upon reasonable request from the authors.

## References

1. Saugstad OD. Reducing global neonatal mortality is possible. Neonatology. 2011;99(4):250–257. doi:10.1159/000320332

2. Murray DM, Bala P, O’Connor CM, Ryan CA, Connolly S, Boylan GB. The predictive value of early neurological examination in neonatal hypoxic–ischaemic encephalopathy and neurodevelopmental outcome at 24 months. Dev Med Child Neurol. 2010;52(2):e55–e59. doi:10.1111/J.1469-8749.2009.03550.X

3. Jacobs SE, Berg M, Hunt R, Tarnow-Mordi WO, Inder TE, Davis PG. Cooling for newborns with hypoxic ischaemic encephalopathy. Cochrane Database of Systematic Reviews. 2013;2013(1). doi:10.1002/14651858.CD003311.pub3

4. Conway JM, Walsh BH, Boylan GB, Murray DM. Mild hypoxic ischaemic encephalopathy and long term neurodevelopmental outcome - A systematic review. Early Hum Dev. 2018;120:80–87. doi:10.1016/J.EARLHUMDEV.2018.02.007

5. Finder M, Boylan GB, Twomey D, Ahearne C, Murray DM, Hallberg B. Two-Year Neurodevelopmental Outcomes after Mild Hypoxic Ischemic Encephalopathy in the Era of Therapeutic Hypothermia. JAMA Pediatr. 2020;174(1):48–55. doi:10.1001/jamapediatrics.2019.4011

6. Törn AE, Hesselman S, Johansen K, Ågren J, Wikström AK, Jonsson M. Outcomes in children after mild neonatal hypoxic ischaemic encephalopathy: A population-based cohort study. BJOG. 2023;130(13):1602–1609. doi:10.1111/1471-0528.17533;WGROUP:STRING:PUBLICATION

7. Chalak L, Latremouille S, Mir I, Sánchez PJ, Sant’Anna G. A review of the conundrum of mild hypoxic-ischemic encephalopathy: Current challenges and moving forward. Early Hum Dev. 2018;120:88–94. doi:10.1016/J.EARLHUMDEV.2018.02.008

8. Lugli L, Guidotti I, Pugliese M, et al. Polygraphic EEG Can Identify Asphyxiated Infants for Therapeutic Hypothermia and Predict Neurodevelopmental Outcomes. Children (Basel). 2022;9(8). doi:10.3390/CHILDREN9081194

9. Cornet MC, Numis AL, Monsell SE, et al. Assessing Early Severity of Hypoxic-Ischemic Encephalopathy: The Role of Electroencephalogram Background in Addition to Sarnat Exam. Journal of Pediatrics. 2025;277. doi:10.1016/j.jpeds.2024.114411

10. Sarnat HB, Sarnat MS. Neonatal Encephalopathy Following Fetal Distress: A Clinical and Electroencephalographic Study. Arch Neurol. 1976;33(10):696–705. doi:10.1001/ARCHNEUR.1976.00500100030012

11. Nyman J, Mikkonen K, Metsäranta M, et al. Poor aEEG background recovery after perinatal hypoxic ischemic encephalopathy predicts postneonatal epilepsy by age 4 years. Clinical Neurophysiology. 2022;143:116–123. doi:10.1016/j.clinph.2022.09.005

12. Jones R, Heep A, Odd D. Biochemical and clinical predictors of hypoxic-ischemic encephalopathy after perinatal asphyxia. J Matern Fetal Neonatal Med. 2018;31(6):791–796. doi:10.1080/14767058.2017.1297790

13. Cornet MC, Numis AL, Wusthoff CJ, et al. Automated EEG Background Analysis and 2-Year Outcomes in Neonatal Hypoxic-Ischemic Encephalopathy. JAMA Netw Open. 2025;8(12):e2548321. doi:10.1001/JAMANETWORKOPEN.2025.48321

14. Lagacé M, Montazeri S, Kamino D, et al. Automated assessment of EEG background for neurodevelopmental prediction in neonatal encephalopathy. Ann Clin Transl Neurol. 2024;11(12):3267–3279. doi:10.1002/ACN3.52233

15. Montazeri S, Nevalainen P, Metsäranta M, Stevenson NJ, Vanhatalo S. Clinical outcome prediction with an automated EEG trend, Brain State of the Newborn, after perinatal asphyxia. Clinical Neurophysiology. 2024;162:68–76. doi:10.1016/j.clinph.2024.03.007

16. Dereymaeker A, Matic V, Vervisch J, et al. Automated EEG background analysis to identify neonates with hypoxic-ischemic encephalopathy treated with hypothermia at risk for adverse outcome: A pilot study. Pediatr Neonatol. 2019;60(1):50–58. doi:10.1016/J.PEDNEO.2018.03.010

17. Kota S, Kang S, Liu YL, et al. Prognostic value of quantitative EEG in early hours of life for neonatal encephalopathy and neurodevelopmental outcomes. Pediatric Research 2024 96:3. 2024;96(3):685–694. doi:10.1038/s41390-024-03255-8

18. Toet MC, Hellström-Westas L, Groenendaal F, Eken P, De Vries LS. Amplitude integrated EEG 3 and 6 hours after birth in full term neonates with hypoxic–ischaemic encephalopathy. Arch Dis Child Fetal Neonatal Ed. 1999;81(1):F19–F23. doi:10.1136/FN.81.1.F19

19. Van Rooij LGM, Toet MC, Osredkar D, Van Huffelen AC, Groenendaal F, De Vries LS. Recovery of amplitude integrated electroencephalographic background patterns within 24 hours of perinatal asphyxia. Arch Dis Child Fetal Neonatal Ed. 2005;90(3):F245. doi:10.1136/ADC.2004.064964

20. Kontio T, Toet MC, Hellström-Westas L, et al. Early neurophysiology and MRI in predicting neurological outcome at 9-10 years after birth asphyxia. Clin Neurophysiol. 2013;124(6):1089–1094. doi:10.1016/J.CLINPH.2012.12.045

21. Weeke LC, Vilan A, Toet MC, Van Haastert IC, De Vries LS, Groenendaal F. A Comparison of the Thompson Encephalopathy Score and Amplitude-Integrated Electroencephalography in Infants with Perinatal Asphyxia and Therapeutic Hypothermia. Neonatology. 2017;112(1):24–29. doi:10.1159/000455819

22. Palva JM, Palva S. Roles of multiscale brain activity fluctuations in shaping the variability and dynamics of psychophysical performance. Prog Brain Res. 2011;193:335–350. doi:10.1016/B978-0-444-53839-0.00022-3

23. Palva S, Palva JM. Functional roles of alpha-band phase synchronization in local and large-scale cortical networks. Front Psychol. 2011;2(SEP). doi:10.3389/FPSYG.2011.00204

24. Palva JM, Monto S, Kulashekhar S, Palva S. Neuronal synchrony reveals working memory networks and predicts individual memory capacity. Proc Natl Acad Sci U S A. 2010;107(16):7580–7585. doi:10.1073/pnas.0913113107

25. Tokariev A, Videman M, Palva MJ, Vanhatalo S. Functional brain connectivity develops rapidly around term age and changes between vigilance states in the human newborn. Cerebral Cortex. 2016;26(12):4540–4550. doi:10.1093/cercor/bhv219

26. Womelsdorf T, Schoffelen JM, Oostenveld R, et al. Modulation of neuronal interactions through neuronal synchronization. Science. 2007;316(5831):1609–1612. doi:10.1126/SCIENCE.1139597

27. Engel AK, Gerloff C, Hilgetag CC, Nolte G. Intrinsic Coupling Modes: Multiscale Interactions in Ongoing Brain Activity. Neuron. 2013;80(4):867–886. doi:10.1016/j.neuron.2013.09.038

28. Yrjölä P, Vanhatalo S, Tokariev A. Neuronal Coupling Modes Show Differential Development in the Early Cortical Activity Networks of Human Newborns. J Neurosci. 2024;44(26). doi:10.1523/JNEUROSCI.1012-23.2024

29. Tokariev A, Vanhatalo S, Palva JM. Analysis of infant cortical synchrony is constrained by the number of recording electrodes and the recording montage. Clinical Neurophysiology. 2016;127(1):310–323. doi:10.1016/J.CLINPH.2015.04.291

30. Tokariev A, Roberts JA, Zalesky A, et al. Large-scale brain modes reorganize between infant sleep states and carry prognostic information for preterms. Nat Commun. 2019;10(1). doi:10.1038/s41467-019-10467-8

31. Tokariev A, Oberlander VC, Videman M, Vanhatalo S. Cortical Cross-Frequency Coupling Is Affected by in utero Exposure to Antidepressant Medication. Front Neurosci. 2022;16. doi:10.3389/fnins.2022.803708

32. Videman M, Tokariev A, Stjerna S, Roivainen R, Gaily E, Vanhatalo S. Effects of prenatal antiepileptic drug exposure on newborn brain activity. Epilepsia. 2016;57(2):252–262. doi:10.1111/epi.13281

33. Videman M, Tokariev A, Saikkonen H, et al. Newborn Brain Function Is Affected by Fetal Exposure to Maternal Serotonin Reuptake Inhibitors. Cerebral Cortex. 2017;27(6):3208–3216. doi:10.1093/cercor/bhw153

34. Yrjölä P, Stjerna S, Palva JM, Vanhatalo S, Tokariev A. Phase-Based Cortical Synchrony Is Affected by Prematurity. Cereb Cortex. 2022;32(10):2265–2276. doi:10.1093/CERCOR/BHAB357

35. Syvälahti T, Tuiskula A, Nevalainen P, et al. Networks of cortical activity show graded responses to perinatal asphyxia. Pediatr Res. 2024;96(1):132–140. doi:10.1038/S41390-023-02978-4

36. McLaren J, Holmes GL, Berg MT. Functional Connectivity in Term Neonates With Hypoxic-Ischemic Encephalopathy Undergoing Therapeutic Hypothermia. Pediatr Neurol. 2019;94:74–79. doi:10.1016/j.pediatrneurol.2019.01.006

37. König S, Tuiskula A, Metsäranta M, et al. Effects of perinatal asphyxia on cortical activity in two-year-old children. Neuroimage Clin. 2026;49:103933. doi:10.1016/J.NICL.2025.103933

38. Sotero RC, Bortel A, Naaman S, et al. Laminar distribution of phase-amplitude coupling of spontaneous current sources and sinks. Front Neurosci. 2015;9(DEC). doi:10.3389/FNINS.2015.00454

39. Hipp JF, Hawellek DJ, Corbetta M, Siegel M, Engel AK. Large-scale cortical correlation structure of spontaneous oscillatory activity. Nat Neurosci. 2012;15(6):884–890. doi:10.1038/NN.3101

40. Azzopardi D v., Strohm B, Edwards AD, et al. Moderate hypothermia to treat perinatal asphyxial encephalopathy. N Engl J Med. 2009;361(14):1349–1358. doi:10.1056/NEJMOA0900854

41. Tuiskula A, Stjerna S, Saure E, Metsäranta M, Haataja L. Perinatal asphyxia with no or mild hypoxic-ischaemic encephalopathy: Two-year neurodevelopmental outcome. Early Hum Dev. 2025;205:106266. doi:10.1016/J.EARLHUMDEV.2025.106266

42. André M, Lamblin MD, d’Allest AM, et al. Electroencephalography in premature and full-term infants. Developmental features and glossary. Neurophysiologie Clinique/Clinical Neurophysiology. 2010;40(2):59–124. doi:10.1016/J.NEUCLI.2010.02.002

43. Vanhatalo S, Matias Palva J, Andersson S, Rivera C, Voipio J, Kaila K. Slow endogenous activity transients and developmental expression of K +-Cl-cotransporter 2 in the immature human cortex. European Journal of Neuroscience. 2005;22(11):2799–2804. doi:10.1111/j.1460-9568.2005.04459.x

44. Vanhatalo S, Palva JM, Holmes MD, Miller JW, Voipio J, Kaila K. Infraslow oscillations modulate excitability and interictal epileptic activity in the human cortex during sleep. Proc Natl Acad Sci U S A. 2004;101(14):5053–5057. doi:10.1073/PNAS.0305375101;PAGE:STRING:ARTICLE/CHAPTER

45. Lachaux JP, Rodriguez E, Martinerie J, Varela FJ. Measuring phase synchrony in brain signals. Hum Brain Mapp. 1999;8(4):194. doi:10.1002/(sici)1097-0193(1999)8:4<194::aid-hbm4>3.0.co;2-c

46. Tokariev A, Stjerna S, Lano A, Metsäranta M, Palva JM, Vanhatalo S. Preterm Birth Changes Networks of Newborn Cortical Activity. Cerebral Cortex. 2019;29(2):814–826. doi:10.1093/cercor/bhy012

47. Odabaee M, Tokariev A, Layeghy S, et al. Neonatal EEG at scalp is focal and implies high skull conductivity in realistic neonatal head models. Neuroimage. 2014;96:73–80. doi:10.1016/J.NEUROIMAGE.2014.04.007

48. Despotovic I, Cherian PJ, De Vos M, et al. Relationship of EEG sources of neonatal seizures to acute perinatal brain lesions seen on MRI: A pilot study. Hum Brain Mapp. 2012;34(10):2402. doi:10.1002/HBM.22076

49. Gramfort A, Papadopoulo T, Olivi E, Clerc M. OpenMEEG: Opensource software for quasistatic bioelectromagnetics. Biomed Eng Online. 2010;9. doi:10.1186/1475-925X-9-45

50. Dale AM, Liu AK, Fischl BR, et al. Dynamic statistical parametric mapping: Combining fMRI and MEG for high-resolution imaging of cortical activity. Neuron. 2000;26(1):55–67. doi:10.1016/S0896-6273(00)81138-1

51. Tadel F, Baillet S, Mosher JC, Pantazis D, Leahy RM. Brainstorm: A User-Friendly Application for MEG/EEG Analysis. Comput Intell Neurosci. 2011;2011(1):879716. doi:10.1155/2011/879716

52. Vinck M, Oostenveld R, Van Wingerden M, Battaglia F, Pennartz CMA. An improved index of phase-synchronization for electrophysiological data in the presence of volume-conduction, noise and sample-size bias. Neuroimage. 2011;55(4):1548–1565. doi:10.1016/J.NEUROIMAGE.2011.01.055

53. Brookes MJ, Woolrich MW, Barnes GR. Measuring functional connectivity in MEG: A multivariate approach insensitive to linear source leakage. Neuroimage. 2012;63(2):910. doi:10.1016/J.NEUROIMAGE.2012.03.048

54. Palva JM, Wang SH, Palva S, et al. Ghost interactions in MEG/EEG source space: A note of caution on inter-areal coupling measures. Neuroimage. 2018;173:632–643. doi:10.1016/J.NEUROIMAGE.2018.02.032

55. Palva S, Palva JM. Discovering oscillatory interaction networks with M/EEG: challenges and breakthroughs. Trends Cogn Sci. 2012;16(4):219–230. doi:10.1016/J.TICS.2012.02.004

56. Asayesh A, Vanhatalo S, Tokariev A. The impact of EEG electrode density on the mapping of cortical activity networks in infants. Neuroimage. 2024;303:120932. doi:10.1016/J.NEUROIMAGE.2024.120932

57. Zalesky A, Fornito A, Bullmore ET. Network-based statistic: identifying differences in brain networks. Neuroimage. 2010;53(4):1197–1207. doi:10.1016/J.NEUROIMAGE.2010.06.041

58. Chalak LF, Pappas A, Tan S, et al. Association Between Increased Seizures During Rewarming After Hypothermia for Neonatal Hypoxic Ischemic Encephalopathy and Abnormal Neurodevelopmental Outcomes at 2-Year Follow-up: A Nested Multisite Cohort Study. JAMA Neurol. 2021;78(12):1484–1493. doi:10.1001/JAMANEUROL.2021.3723

59. Meder U, Cseko AJ, Szakacs L, et al. Longitudinal Analysis of Amplitude-Integrated Electroencephalography for Outcome Prediction in Hypoxic-Ischemic Encephalopathy. Journal of Pediatrics. 2022;246:19–25.e5. doi:10.1016/j.jpeds.2022.04.013

60. De Wispelaere LA, Ouwehand S, Olsthoorn M, et al. Electroencephalography and brain magnetic resonance imaging in asphyxia comparing cooled and non-cooled infants. European Journal of Paediatric Neurology. 2019;23(1):181–190. doi:10.1016/j.ejpn.2018.09.001

61. Ferrari F, Bondi C, Lugli L, et al. The Dammiss EEG Score: A New System to Quantify EEG Abnormalities and Predict the Outcome in Asphyxiated Newborns. J Clin Med. 2025;14(6). doi:10.3390/JCM14061920

62. Langeslag JF, Onland W, Groenendaal F, et al. Association Between Seizures and Neurodevelopmental Outcome at Two and Five Years in Asphyxiated Newborns With Therapeutic Hypothermia. Pediatr Neurol. 2024;153:152–158. doi:10.1016/j.pediatrneurol.2024.01.023

63. Wang X, Liu H, Kota S, et al. EEG phase-amplitude coupling to stratify encephalopathy severity in the developing brain. Comput Methods Programs Biomed. 2022;214:106593. doi:10.1016/J.CMPB.2021.106593

64. Wang X, Liu H, Ortigoza EB, et al. Feasibility of EEG Phase-Amplitude Coupling to Stratify Encephalopathy Severity in Neonatal HIE Using Short Time Window. Brain Sci. 2022;12(7). doi:10.3390/BRAINSCI12070854

65. Koolen N, Dereymaeker A, Räsänen O, et al. Interhemispheric synchrony in the neonatal EEG revisited: Activation synchrony index as a promising classifier. Front Hum Neurosci. 2014;8(DEC):1030. doi:10.3389/FNHUM.2014.01030/ABSTRACT

66. Tuiskula A, Metsäranta M, Toiviainen-Salo S, Vanhatalo S, Haataja L. Profile of minor neurological findings after perinatal asphyxia. Acta Paediatr. 2022;111(2):291–299. doi:10.1111/APA.16133

67. Wang X, Liu H, Ortigoza EB, et al. Feasibility of EEG Phase-Amplitude Coupling to Stratify Encephalopathy Severity in Neonatal HIE Using Short Time Window. Brain Sci. 2022;12(7):854. doi:10.3390/brainsci12070854

68. Shankaran S, Pappas A, McDonald SA, et al. Childhood Outcomes after Hypothermia for Neonatal Encephalopathy. Published online 2012. doi:10.1056/NEJMoa1112066

69. Mwaniki MK, Atieno M, Lawn JE, Newton CRJC. Long-term neurodevelopmental outcomes after intrauterine and neonatal insults: a systematic review. Lancet. 2012;379(9814):445–452. doi:10.1016/S0140-6736(11)61577-8

70. Grossmann K, Westblad M, … MBA of D in, 2023 undefined. Outcome at early school age and adolescence after hypothermia-treated hypoxic–ischaemic encephalopathy: an observational, population-based study. fn.bmj.comKR Grossmann, ME Westblad, M Blennow, K LindströmArchives of Disease in Childhood-Fetal and Neonatal Edition, 2023•fn.bmj.com. Accessed April 21, 2026. https://fn.bmj.com/content/108/3/295?ref=the-incubator.org

71. Marlow N, Rose A, Rands C, in EDA of D, 2005 undefined. Neuropsychological and educational problems at school age associated with neonatal encephalopathy. fn.bmj.comN Marlow, AS Rose, CE Rands, ES DraperArchives of Disease in Childhood-Fetal and Neonatal Edition, 2005•fn.bmj.com. Accessed April 21, 2026. https://fn.bmj.com/content/90/5/F380?fbclid=IwAR1So154uX7xdltQVpTAN11wUiUYOzvYdW8MCwj5CZaxMwBBjJ8fh343Oi0

72. Lee-Kelland R, Jary S, Tonks J, … FCA of D in, 2020 undefined. School-age outcomes of children without cerebral palsy cooled for neonatal hypoxic–ischaemic encephalopathy in 2008–2010. fn.bmj.comR Lee-Kelland, S Jary, J Tonks, FM Cowan, M Thoresen, E ChakkarapaniArchives of Disease in Childhood-Fetal and Neonatal Edition, 2020•fn.bmj.com. Accessed April 21, 2026. https://fn.bmj.com/content/105/1/8.abstract

