## Supplementary Material for "Neonatal EEG network activity associates with 2-year neurodevelopment after perinatal asphyxia"

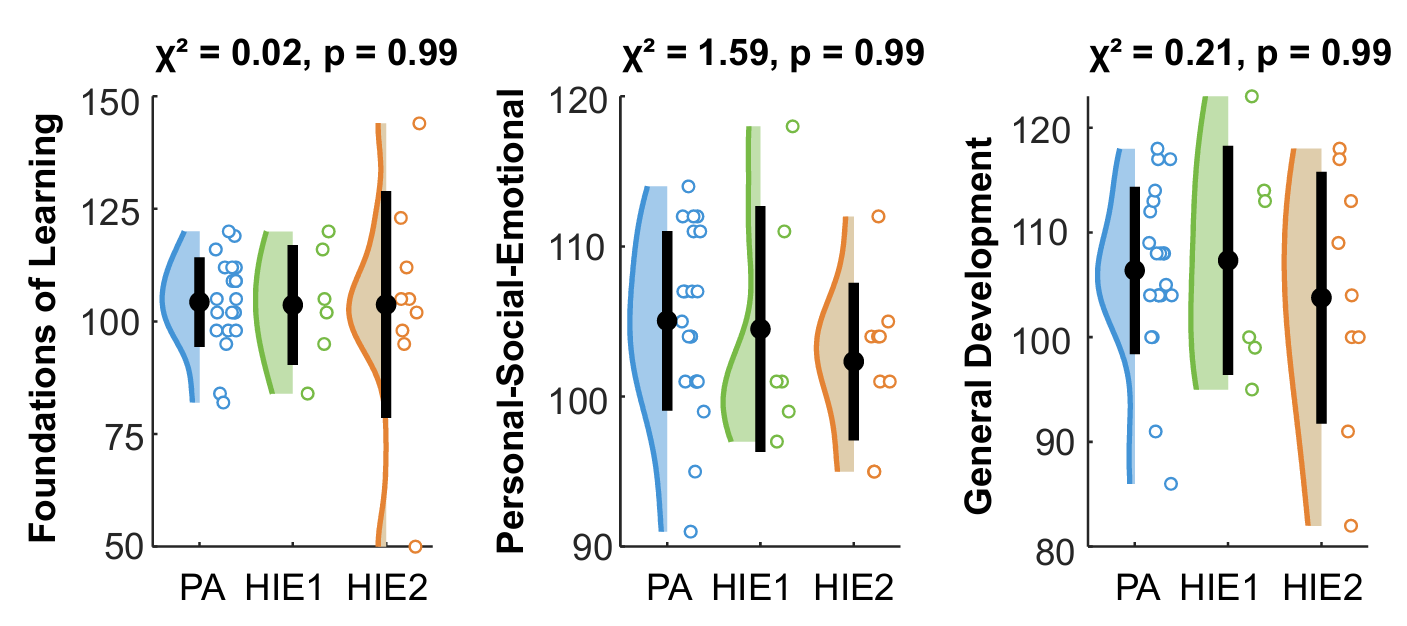

**Supplementary Figure 1. GMDS-III scores across HIE severity groups.** Violin plots show the distribution of 2-year GMDS-III Foundations of Learning, Personal-Social-Emotional, and General Development scores across clinical severity groups. Black bars indicate the interquartile range (IQR), and black dots show the median. χ² values are from the Kruskal–Wallis test, and displayed p-values are FDR-corrected. No significant differences were observed between the groups. PA = perinatal asphyxia without HIE, HIE1 = mild hypoxic-ischemic encephalopathy, HIE2 = moderate hypoxic-ischemic encephalopathy.

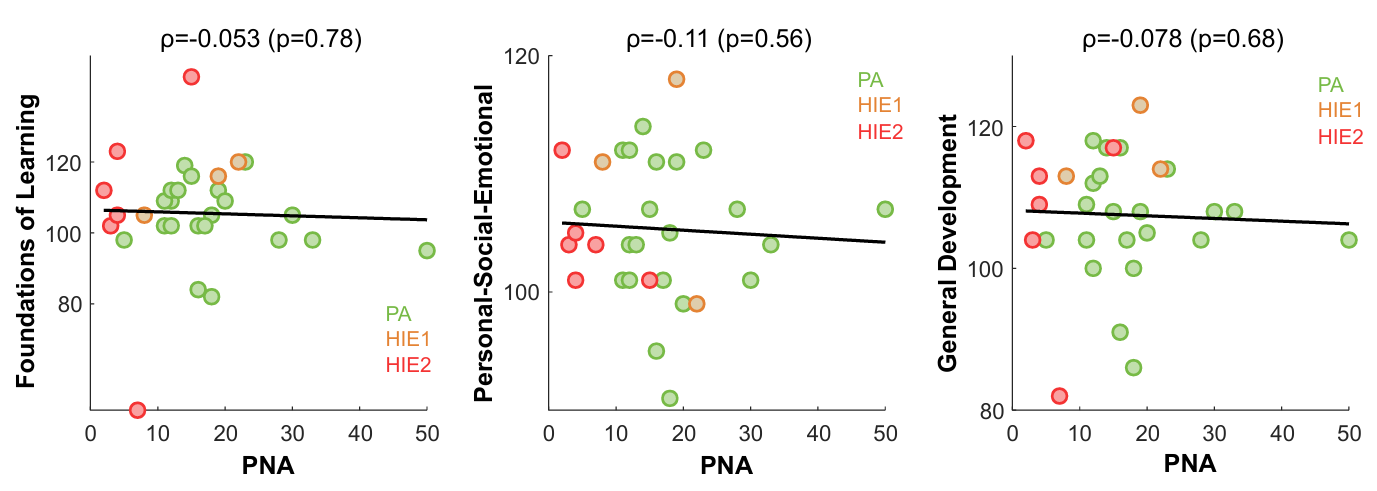

**Supplementary Figure 2. Correlation between postnatal age at EEG recording and 2-year GMDS-III outcomes.** Scatterplots show the relationships between postnatal age at EEG recording (PNA) and 2-year GMDS-III Foundations of Learning, Personal-Social-Emotional, and General Development scores. Spearman correlation coefficients (ρ) with FDR-corrected p-values are shown above each panel. No significant associations were found. The colored dots represent individual infants. Green = perinatal asphyxia without HIE (PA), yellow = mild HIE (HIE1), red = moderate HIE (HIE2).

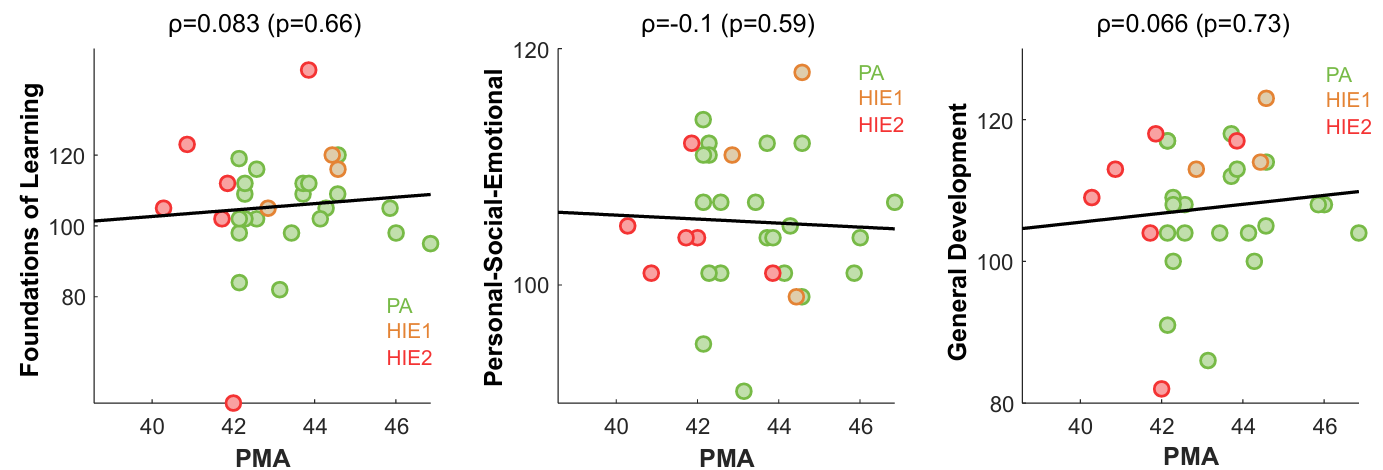

**Supplementary Figure 3. Correlation between postmenstrual age at EEG recording and 2-year GMDS-III outcomes.** Scatterplots show the relationships between postmenstrual age at EEG recording (PNA) and 2-year GMDS-III Foundations of Learning, Personal-Social-Emotional, and General Development scores. Spearman correlation coefficients (ρ) with FDR-corrected p-values are shown above each panel. No significant associations were found. The colored dots represent individual infants. Green = perinatal asphyxia without HIE (PA), yellow = mild HIE (HIE1), red = moderate HIE (HIE2).

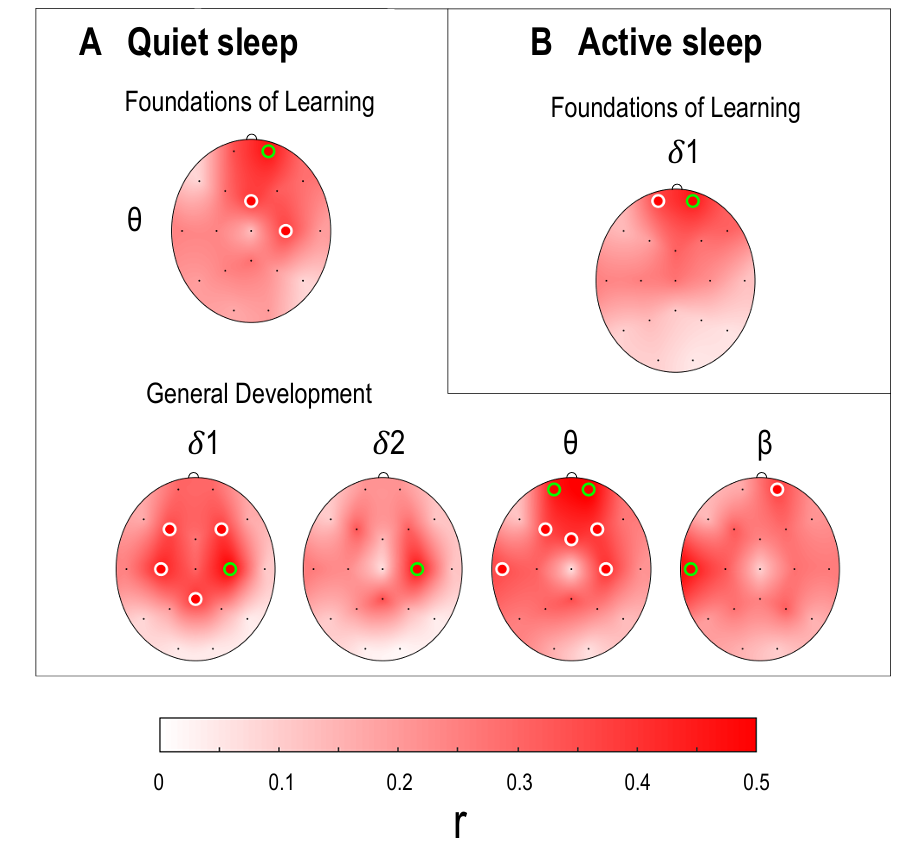

**Supplementary Figure 4. Associations between local EEG amplitudes and 2-year neurodevelopment after adjustment for postmenstrual age.** Scalp colormaps show associations between local EEG amplitudes and 2-year GMDS-III Foundations of Learning and General Development scores in quiet sleep (A) and active sleep (B). Frequency bands are shown above each plot (𝛿1: low delta, 𝛿2: high delta, θ: theta, β: beta). Color indicates the strength of association, expressed as partial correlation (r) derived from the regression model. Circles indicate electrodes where amplitudes showed significant associations with GMDS-III scores (p < 0.05), while green circles denote electrodes that also passed FDR correction for multiple comparisons.

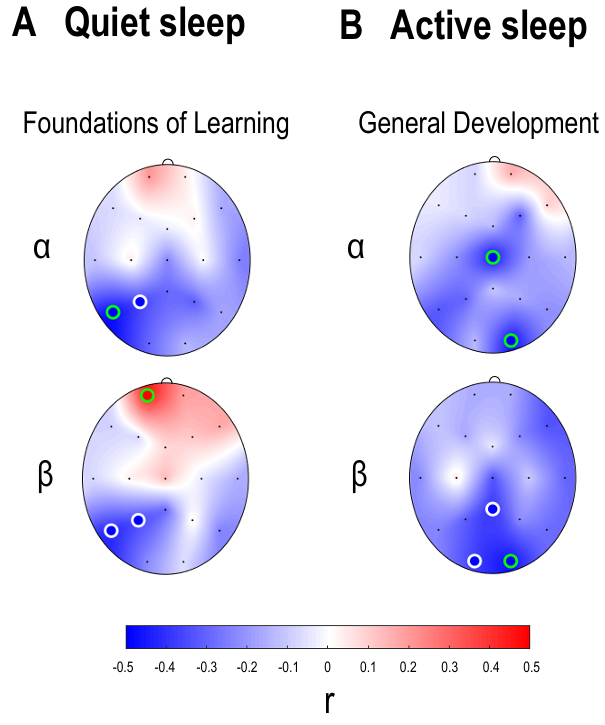

**Supplementary Figure 5. Associations between local phase-amplitude coupling (PAC) and 2-year neurodevelopment after adjustment for postmenstrual age.** Scalp colormaps show associations between local PAC and 2-year GMDS-III Foundations of Learning and General Development scores in quiet sleep (A) and active sleep (B). Frequency bands are shown next to each plot (α: alpha, β: beta). Color indicates the strength of association, expressed as partial correlation (r) derived from the regression model. Circles mark electrodes at which PAC was significantly associated with the corresponding GMDS-III scores (p < 0.05), with green circles indicating electrodes that also survived FDR correction across electrodes.

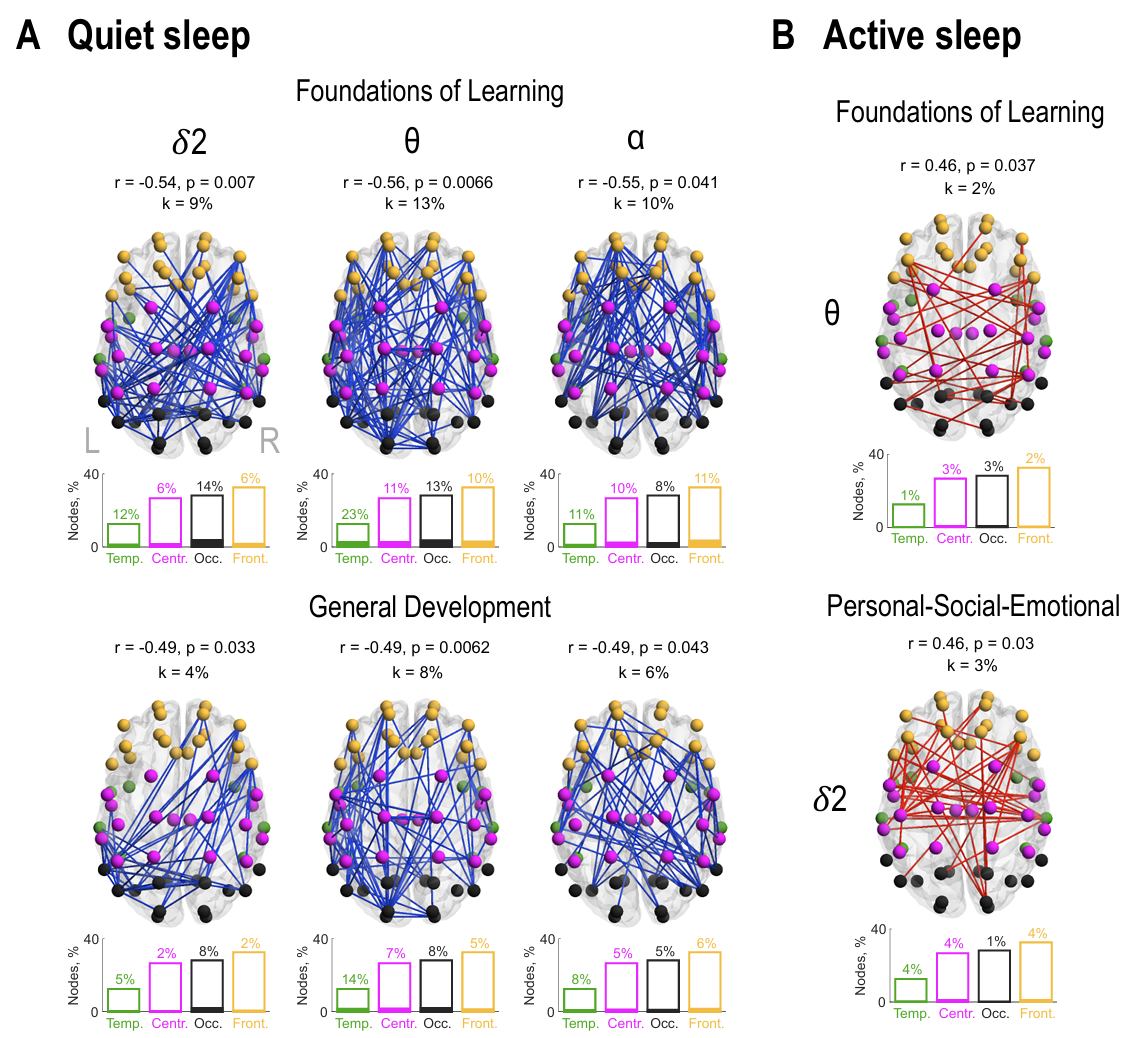

**Supplementary Figure 6. Cortical PPC networks association with 2-year outcomes with adjustment for postmenstrual age.** Network plots show associations between large-scale phase–phase correlations (PPC) and 2-year GMDS-III outcomes in quiet sleep (A) and active sleep (B). Frequency bands shown above or next to each plot (𝛿2: high delta, θ: theta, α: alpha). Blue networks indicate negative associations and red indicate positive associations. Node colors highlight distinct anatomical regions: temporal = green, central = magenta, occipital = black, and frontal = orange. Bar height indicates the maximum possible involvement of nodes from each anatomical region, while the filled portion shows their observed involvement in specific networks. Partial correlation coefficients (r), derived from the regression models, corresponding family-wise error-corrected p-values, and network densities (k) are shown above each network.

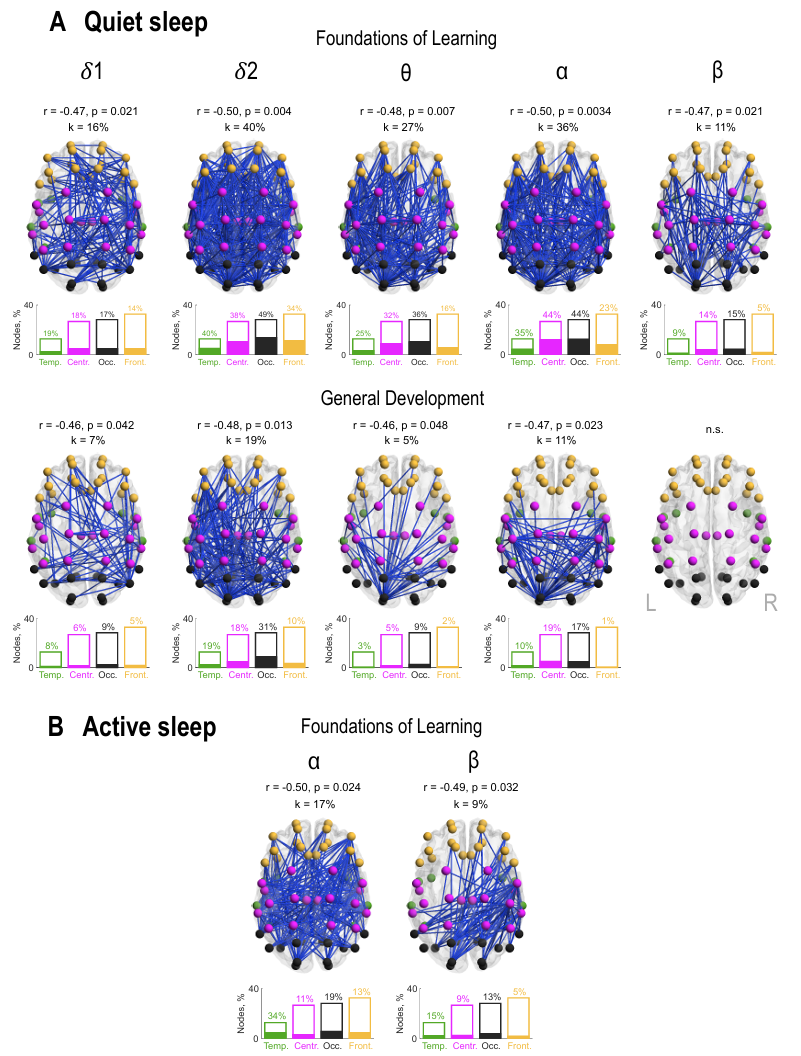

**Supplementary Figure 7. Cortical AAC networks association with 2-year outcomes with adjustment for postmenstrual age.** Network plots show associations between large-scale amplitude–amplitude correlations (AAC) and 2-year GMDS-III outcomes in quiet sleep (A) and active sleep (B). Frequency bands are shown above plots (𝛿1: low delta, 𝛿2: high delta, θ: theta, α: alpha, β: beta). Node colors indicate anatomical regions (temporal = green, central = magenta, occipital = black, and frontal = orange), and the bar plots below each network show the involvement of specific anatomical regions in the association network, indicated by the filled portions of the bars. Partial correlation coefficients (r), derived from the regression models, corresponding family-wise error-corrected p-values, and network densities (k) are shown above each network. N.s.: no significant association.
